# Development and validation of a tool to stratify the treatment effect of low-dose aspirin in patients with cardiovascular disease: VISTA (Vascular Intervention Stratification Tool for Aspirin)

**DOI:** 10.1101/2024.06.07.24308636

**Authors:** Yekai Zhou, Joseph Edgar Blais, Esther Wai Yin Chan, Tak-Wah Lam, Kai Hang Yiu, Hung Fat Tse, Celine SL Chui, Ruibang Luo

## Abstract

**Background:** Aspirin resistance, as determined by measuring platelet aggregation function, has been investigated as a proxy outcome for clinical treatment failure with low-dose aspirin (occurrence of cardiovascular events despite regular aspirin intake). However, among adults with cardiovascular disease (CVD), there is no method to predict aspirin treatment failure using routinely available clinical data. We aimed to develop and internally validate the Vascular Intervention Stratification Tool for Aspirin (VISTA), a model that predicts inter-individual variability in clinical response to low-dose aspirin.

**Methods:** We used electronic health records of the Hong Kong Hospital Authority to identify derivation (n=48,743) and validation (n=322,731) cohorts consisting of individuals diagnosed with CVD between January 1, 2015 to December 31, 2018. The composite outcome of recurrent CVD event included the diagnosis of coronary heart disease, ischemic stroke, and peripheral artery disease after low-dose aspirin initiation (≤ 100 mg). Cox proportional hazards regression with the least absolute shrinkage and selection operator regularization was used to identify the most strongly associated and relevant risk factors for aspirin treatment failure. One-year hazard ratio (HR) was estimated across different risk categories for aspirin treated vs. untreated individuals.

**Results:** The derivation cohort included 1,623 individuals who initiated low-dose aspirin after their CVD diagnosis. Among 109 variables available, six were selected as model inputs: atrial fibrillation, dyslipidemia, hyperglycemia, polypharmacy, neutrophilia, and elevated serum creatine kinase. In the model validation cohort, we identified 22,192 individuals who initiated low-dose aspirin and 3,747 individuals without aspirin, other antiplatelets, or anticoagulants. Results of the model validation demonstrated a strong graded association between the number of VISTA risk factors and the one-year risk of CVD. Compared to untreated individuals, low-dose aspirin use with no VISTA risk factors had the lowest HR for CVD (0.68, 95% CI of 0.57 to 0.81). For low-dose aspirin user with 1-2 VISTA risk factors, HRs was 0.87 (0.81 to 0.93). The presence of 3-6 VISTA risk factors was associated with aspirin treatment failure (HR 0.99; 95% CI of 0.88 to 1.12), which occurred in approximately 20% of patients in our validation cohort.

**Conclusions:** VISTA can predict the heterogeneity of low-dose aspirin’s treatment effect against recurrent CVD. VISTA could be used to stratify patients based on six readily available risk factors and inform patients and clinicians about the potential benefits of aspirin therapy and the potential for alternate antiplatelet treatments.

**Clinical perspective:** *What is new?:*

- We developed VISTA (Vascular Intervention Stratification Tool for Aspirin), the first tool that enables the stratification of low-dose aspirin treatment effect for the secondary prevention of cardiovascular disease (CVD). By utilizing six easily accessible clinical risk factors (atrial fibrillation, dyslipidemia, hyperglycemia, polypharmacy, neutrophilia, and elevated serum creatine kinase), VISTA allows for the assessment of aspirin suitability before prescription.
- VISTA can differentiate patients who are likely to benefit significantly from aspirin treatment (0 risk factors, hazard ratio [HR] of 0.68) from those who may experience aspirin treatment failure (3-6 risk factors, HR of 0.99).

*What are the clinical implications?:*

- Estimated from a high-quality contemporary validation cohort, there are approximately 20% of patients with CVD who may experience aspirin treatment failure.
- By utilizing VISTA, healthcare providers can personalize aspirin treatment, optimizing its effectiveness and minimizing the potential for treatment failure. This tool empowers clinicians to make more accurate and tailored decisions in prescribing low-dose aspirin for secondary prevention of cardiovascular disease, ultimately leading to improved patient outcomes.

## Introduction

Cardiovascular diseases (CVD), such as coronary heart disease and stroke, are the leading cause of death worldwide, resulting in an estimated 18.6 million fatalities in 2019.^1,2^ Aspirin, or acetylsalicylic acid, has long been recognized as a crucial intervention to improve CVD outcomes due to its antiplatelet properties, particularly in secondary prevention of CVD.^3,4^ However, the occurrence of occlusive CVD events despite regular aspirin intake, a phenomenon known as aspirin treatment failure, poses a significant clinical challenge.^5–10^ Identifying patients who may have limited benefit from CVD prevention with aspirin but are exposed to an elevated risk of major organ bleeding is essential for appropriate aspirin prescription and selection of suitable antiplatelet treatments for CVD prevention.^11^ Although methods exist to indirectly query aspirin effectiveness via platelet aggregation function tests, there are currently no solutions available to predict aspirin treatment failure in clinical settings.^5,12–15^

Existing methods for evaluating aspirin effectiveness involve assays that test a laboratory phenomenon called aspirin resistance.^5^ These assays, including the platelet function analyzer (PFA-100), whole blood aggregometry (WBA), and VerifyNow aspirin assay (VerifyNow), quantify and compare the platelet aggregation function before and after the administration of aspirin.^5,12–15^ However, these assays have limitations when it comes to aspirin prescriptions. First, they serve only as risk indicators, similar to age and lipid levels; they cannot measure the treatment effect of aspirin.^16,17^ Second, these assays utilize different techniques, resulting in poor agreement even among the same group of patients.^18–25^ This lack of consistency adds to the difficulty for clinicians in selecting the most appropriate assay. Third, these assays are not endorsed by clinical guidelines and are not typically available in general hospital settings, limiting their use as routine tests for aspirin prescription reference.^26,27^ Therefore, there is an urgent need for a method based on easily accessible patient characteristics that can directly estimate the treatment effect of aspirin in CVD prevention.

In this study, we present the development and validation of the Vascular Intervention Stratification Tool for Aspirin (VISTA), which was designed to stratify the treatment effect of aspirin for the prevention of recurrent CVD events. VISTA is based on six easily accessible patient characteristics that can be assessed prior to aspirin prescription in actual clinical settings. We conducted a large, comprehensive cohort study in Hong Kong, encompassing a wide range of potential risk factors. By employing advanced big data approaches, we analyzed this extensive dataset to identify the most significant risk factors contributing to the variation in aspirin treatment effects. The validity of the identified risk factors was then confirmed in a separate validation cohort, which demonstrated their accuracy in stratifying the treatment effects of aspirin. Our method provides a straightforward estimation of aspirin’s treatment effect, serving as a tool to assess its potential benefits for individual patients. Leveraging VISTA’s capabilities, clinicians gain early access to this information, enabling informed decisions about aspirin therapy suitability.

## Methods

### Data source

Two contemporary cohorts were identified for model derivation and validation from the electronic health records of the Hong Kong Hospital Authority (HA) with rigorous inclusion criteria (Figure 1). The HA is a statutory body and the largest public healthcare provider of Hong Kong. It provides government subsidized primary, secondary, and tertiary care for all residents who want it, capturing over 70% of all hospitalizations in Hong Kong.^28^ Previous studies demonstrated the high validity of the data source, with a positive predictive value of 85% for myocardial infarction (MI) and 91% for stroke.^29^ Because patients are provided outpatient medications at very low out-of-pocket cost, dispensing records for low-dose aspirin are recorded in the HA database. This makes it suitable to investigate the chronic effects of low-dose aspirin, which may not be feasible in many other health systems where patients purchase aspirin as an over-the-counter medication without prescription or recording.^30^

**Figure 1.**
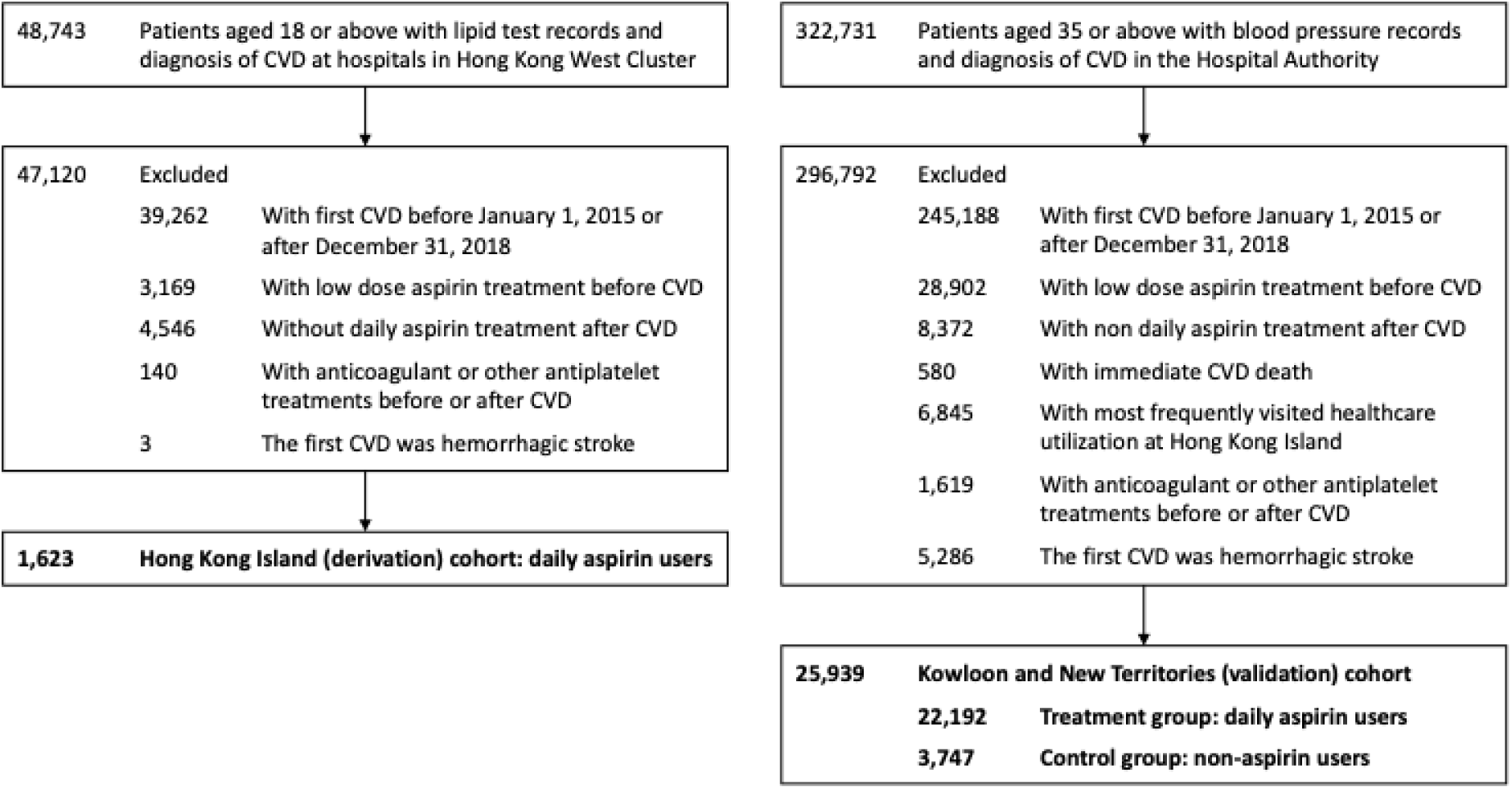
**Selection of patients.** Hong Kong West Cluster is a part of Hong Kong Island. CVD = cardiovascular diseases.

### Study cohorts and eligibility criteria

We included patients who 1) had their first CVD diagnosis recorded between 1 January 2015 and 31 December 2018, 2) did not have an aspirin prescription dispensed before the index CVD event, and 3) had a prescription record for low-dose (≤ 100mg) aspirin daily until the end of study follow-up, or did not have any prescription record for aspirin at any dose until the end point of their study (as the control group to compare with the treated patients to calculate the treatment effect of aspirin).The cohort entry date was the date of their first diagnosis of CVD in any inpatient or outpatient setting. Patients were followed until the earliest occurrence of the composite recurrent CVD outcome, date of registered death, one year after their index CVD diagnosis, or the study end date (31 December 2019). We did not include patients with CVD diagnosis within one year before the study end date to ensure the patients would have sufficient exposure to aspirin therapy. Daily aspirin users whose most frequently visited healthcare facility within the study period was on Hong Kong Island were categorized as the Hong Kong Island cohort for model derivation. Patients with their most frequently visited healthcare facility within the study period was on Kowloon and New Territories were categorized as the Kowloon and New Territories cohort for model validation. The daily aspirin users were in the treatment group and the non-aspirin users in the untreated (control) group.

### Exposure, outcomes and risk factor variables

The included patients had a CVD diagnosis (coronary heart disease, ischemic stroke, and peripheral artery disease) between January 1, 2015, and December 31, 2018. Patients were not prescribed low-dose (≤ 100 mg) aspirin before the CVD event. The control group patients were not treated with low-dose aspirin before their censored date. The control group patients received sufficient daily low-dose aspirin treatment from the date of CVD event to their censored date.

The outcome was the diagnosis of CVD defined by the International Classification of Diseases, Ninth Revision, Clinical Modification (ICD-9-CM) codes. The outcome was a composite of coronary heart disease, ischaemic or haemorrhagic stroke, and peripheral artery disease (Supplementary Table 1).

We compiled an extensive list of over 100 variables to ensure the selection of representative risk factors. The list encompassed clinical laboratory tests, disease and medication history, family history of disease, demographic factors, and healthcare utilization (Table 2). Diagnoses and procedures were defined by ICD-9-CM codes (Supplementary Table 2), and medication history was defined by the British National Formulary (BNF) sections (Supplementary Table 3).

### Statistical analysis

A robust feature selection pipeline was applied to identify the risk variables for model derivation.^31,32^ Multivariate imputation with chained equations (MICE) was used to generate one imputed dataset to replace the missing values of clinical laboratory tests.^33^ MICE is a principled method for dealing with missing data and is extremely reliable on high-dimensional datasets with various missing patterns.^34^ For better statistical reliability and clinical utility, risk variables with missing rates below 20% (e.g., clinical laboratory tests) and an event rate above 1% (e.g., disease and medication history) were passed for feature selection. We employed a Cox proportional hazards model (CPH) with the least absolute shrinkage and selection operator (LASSO) regularization to shortlist statistically significant (p value<0.05) risk variables.^35,36^ CPH is the most widely used multivariate statistical model for survival analysis.^37,38^ Its regression coefficients can be interpreted as hazard ratios which can be easily understood by clinicians for better decision-making. LASSO is a robust feature selection method. It selects the most representative but independent set of risk variables, which is reliable when downstream manual prioritization is required. The final list of risk variables was also determined based on current clinical evidence to ensure the final set of risk variables are comprehensive and relevant to CVD prognosis. The identified continuous risk variables were converted to categorical form based on clinical diagnosis criteria. Each risk variable was weighted as one point, and the risk category was defined as the total number of risk factors for each patient.

Patients in the validation cohort were assigned risk categories based on the identified risk variables. One-year CVD risk was calculated for patients within different risk categories in the treatment and control groups respectively using the Kaplan-Meier method.^39^ The treatment effect and the confidence interval across different risk categories were calculated as the coefficient in CPH, which was trained on the composite of treatment and control group patients with treatment as a covariate in each risk category. All analyses were conducted using Python (version 3.9.1), with its add-on package lifelines.^40^ This study report is in accordance with the TRIPOD statement.^41^ Ethical approval for this study was granted by the Institutional Review Board of The University of Hong Kong/HA Hong Kong West Cluster (UW24-124).

## Results

### Study cohorts

For the derivation cohort, we initially identified 48,743 patients aged 18 or above with lipid test records and diagnosis of CVD at hospitals in the Hong Kong West Cluster. We excluded 39,262 patients whose first CVD event occurred before January 1, 2015, or after December 31, 2018. We excluded 3,169 patients with low dose aspirin treatment before their first CVD. We excluded 4,546 patients with no daily aspirin treatment after CVD. We excluded 140 patients with anticoagulant or other antiplatelet treatments before or after CVD. We excluded 3 patients with the first CVD diagnosis of hemorrhagic stroke. Overall, 1,623 patients (selected out of 48,743 patients) who took aspirin daily were included in the Hong Kong Island cohort for model derivation.

For the validation cohort, we initially identified a cohort of 322,731 patients aged 35 or above with blood pressure records and diagnosis of CVD by the Hospital Authority. Similar exclusion criteria were applied to the validation cohort. A total of 245,188 patients were excluded who had their first CVD event before January 1, 2015 or after December 31, 2018. A total of 28,902 of patients were excluded who had low dose aspirin treatment before their first CVD. A total of 8,372 patients were excluded who had received aspirin treatment after CVD, but not daily. A total of 580 patients were excluded because of immediate CVD death. To ensure no overlap with the derivation cohort, we excluded 6,845 patients with most frequently visited healthcare utilization on the Hong Kong Island. We excluded 1,619 patients with anticoagulant or other antiplatelet treatments before or after CVD. We excluded 5,286 patients with the first CVD diagnosis of hemorrhagic stroke. Overall, 25,939 patients (selected out of 322,731 patients) were included in the Kowloon and New Territories cohort for model validation; 22,192 daily aspirin users were assigned to the treatment group and 3,747 non aspirin users were assigned to the control group.

### Baseline characteristics

The baseline characteristics were similar across the cohorts and subgroups, to ensure the reliability of our selection criteria and support the subsequent model derivation and validation (Table 1). The median age at the cohort entry date was 67 to 75, and 35–48% of the patients were female. A considerable proportion of the patients had comorbidities: 12-23% with dyslipidemia, 30-40% with hypertension, 19-20% with diabetes, 3-10% with atrial fibrillation, 3-5% with chronic kidney disease, and 6-7% with congestive heart failure. Regarding medication history, 18-21% of patients had received statin therapy, 15-22% of patients had taken antidiabetic drugs, and 42-65% of patients were on antihypertensive drugs. The median number of concurrent medications was 6-7.

**Table 1.** Baseline characteristics of the derivation and validation cohorts.

|  | Derivation cohort:<br>Hong Kong Island<br>(n=1,623) |  | Validation cohort: Kowloon and New Territories |  |  |  |
| --- | --- | --- | --- | --- | --- | --- |
|  |  |  | Treatment group<br>(n=22,192) |  | Control group<br>(n=3,747) |  |
| Demographic factors |  |  |  |  |  |  |
| Age | 67 | (58-78) | 68 | (59-79) | 75 | (63-85) |
| Female sex | 561 | (35%) | 8,404 | (38%) | 1,807 | (48%) |
| Male sex | 1,062 | (65%) | 13,788 | (62%) | 1,940 | (52%) |
| Disease history |  |  |  |  |  |  |
| Atrial fibrillation | 74 | (5%) | 708 | (3%) | 390 | (10%) |
| Dyslipidemia | 215 | (13%) | 5,180 | (23%) | 431 | (12%) |
| Diabetes | 315 | (19%) | 4,404 | (20%) | 725 | (19%) |
| Hypertension | 492 | (30%) | 7,694 | (35%) | 1,488 | (40%) |
| Chronic kidney disease | 43 | (3%) | 563 | (3%) | 179 | (5%) |
| Congestive heart failure | 109 | (7%) | 1,379 | (6%) | 274 | (7%) |
| Medication history |  |  |  |  |  |  |
| Statins | 288 | (18%) | 4,638 | (21%) | 784 | (21%) |
| Antidiabetic drugs | 242 | (15%) | 4,487 | (20%) | 841 | (22%) |
| Antihypertensive | 680 | (42%) | 12,556 | (57%) | 2453 | (65%) |
| Count of medication | 6 | (4-8) | 6 | (5-9) | 7 | (5-10) |
| Clinical laboratory tests |  |  |  |  |  |  |
| Neutrophil, 10^9/L | 4.8 | (3.6-6.7, 0%) | 5.4 | (4.0-7.7, 1%) | 6.2 | (4.3-9.8, 1%) |
| Creatine kinase, IU/L | 140 | (83-319, 14%) | 136 | (82-304, 10%) | 112 | (63-219, 11%) |
| Glucose (fasting), mmol/L | 5.7 | (5.2-6.9, 1%) | 5.8 | (5.2-7.0, 1%) | 5.7 | (5.1-7.0, 9%) |

All data in n (%), median (interquartile range), or median (interquartile range, proportion of missing data).

### Model development

The complete list of all available variables and the details of the feature selection process are shown in Table 2. There were initially 109 risk variables under consideration. We excluded 38 of them with a high missing rate (>20%) or low event rate (<1%) from subsequent LASSO regression for better statistical reliability and clinical utility. The LASSO regression on the remaining variables identified 7 statistically significant (p value < 0.05) variables. We shortlisted 4 of them based on clinical evidence. We replaced the use of antiplatelet drugs with fasting glucose level as a measure of diabetic inclination due to the higher data completeness of the latter. We added atrial fibrillation to the final list considering its established role in suboptimal aspirin effectiveness.^42,43^ We translated the selected numerical variables, namely neutrophil count, creatine kinase, glucose levels, and concurrent medication, into categorical variables based on specific diagnostic criteria. Neutrophil count was categorized as neutrophilia (neutrophil count > 7.5 * 10^9/L), creatine kinase was categorized as elevated CK (defined as CK > 400 IU/L), glucose levels were categorized as hyperglycemia (glucose > 7 mmol/L), and concurrent medication was categorized as polypharmacy (concurrent medication > 4).

**Table 2.** Summary of included 109 variables assessed in the model derivation cohort.

| Categories (number of variables) | Risk variables |
| --- | --- |
| **Demographic factors (2)** | age, sex |
| **Family history of disease (2)** | diabetes, ~~cardiovascular disease~~ |
| **Healthcare utilization (3)** | outpatient visits per year*, accident and emergency visits per year, inpatient visits per year |
| **Clinical laboratory tests (39)** | neutrophil*^, creatine kinase (total)*^, monocyte*, glucose (fasting)^, aspartate transaminase, alanine aminotransferase, low-density lipoprotein cholesterol, hemoglobin A1c, prothrombin time, potassium (serum), estimated glomerular filtration rate, triglycerides, basophil, albumin, international normalized ratio, bilirubin (total), total cholesterol, lymphocyte, creatinine (serum), platelet, red blood cell, high-density lipoprotein cholesterol, body mass index, calcium (serum), white blood cell, alkaline phosphatase, sodium (serum), eosinophil, hemoglobin, ~~thyroid stimulating hormone, diastolic blood pressure, systolic blood pressure, erythrocyte sedimentation rate, C-reactive protein, bicarbonate (serum), blood pH, free thyroxine, arterial partial pressure of oxygen, troponin-I~~ |
| **Medication history (24)** | antidiabetic drugs*, count of medication*^, statins, antihypertensive drugs, non-steroidal anti-inflammatory drugs, corticosteroids, proton-pump inhibitors, H2-receptor antagonists, nicotine replacement therapy, antiarrhythmic drugs, antithyroid drugs, psychotropic drugs, nitrates, thyroid hormones, fibrates, other non-statin lipid-modifying drugs, ~~cardiac glycosides, omega-3 fatty acids, PCSK9 inhibitors, cholesterol absorption inhibitors, bile acid sequestrants, testosterone, oestrogen, niacin~~ |
| **Disease history (39)** | dyslipidemia*^, pacemaker implantation, atrial fibrillation^, hypertension, diabetes, congestive heart failure, thyroid disease, arrhythmia and conduction disorders, obesity, oxygen therapy/ventilator/intubation, asthma, injury and poisoning, severe mental illness, dementia, liver disease, chronic obstructive pulmonary disease, cancer, renal disease, chronic kidney disease, dialysis, ~~Huntington's disease, erectile dysfunction, systemic lupus erythematosus, Parkinson's disease, nephrotic syndrome, Creutzfeldt-Jakob disease, Down's syndrome, defibrillator insertion, smoker, alcohol user, heart transplantation, rheumatoid arthritis, hypothyroidism, cardioversion, muscle pain or myopathy or rhabdomyolysis, migraine, cardiac wall/valve/shunt replacement/repairment, cardiomyopathy, major organ bleeding~~ |

Strikethrough: excluded from LASSO regression due to high missing rate (>20%) or low event rate (<1%).
\*Significant variables (p-value<0.05) in LASSO regression. ^Selected variables for final modeling. PCSK9 = Proprotein convertase subtilisin/kexin type 9.

The identified risk factors encompassed a comprehensive spectrum of direct and indirect causes that contributed aspirin suboptimal effects, which were supported by the literature. First, while aspirin is more effective in atherosclerotic CVD, its effectiveness in other types of CVD may be limited.^44^ Aspirin has limited efficacy for stroke prevention in patients with atrial fibrillation.^43^ Neutrophilia may serve as an indicator of inflammation signaling arteritis which compromise the efficacy of aspirin.^44,45^ Elevated CK, an indicator of muscle damage, is a possible sign of cardiomyopathy or myonecrosis, which could potentially impact the response to aspirin treatment.^46,47^ Specific comorbidities and prevalent drug use may also affect the treatment effect of aspirin. Polypharmacy can lead to drug interactions that interfere with the effect of aspirin. Hyperglycemia and dyslipidemia may hinder the intended antiplatelet effects of aspirin by increasing the production of prostaglandin F2-like compounds.^48–50^ The final list of the 6 risk factors was then utilized in the CPH. The parameter estimates for each of the 6 predictor variables showed statistical significance and were of similar magnitude (Table 3).

**Table 3.** Multivariate stratification model for aspirin treatment effect.

| Variable | HR (95%CI) | p-value | points |
| --- | --- | --- | --- |
| Atrial fibrillation | 1.73 (1.33-2.24) | <0.05 | 1 |
| Dyslipidemia | 1.19 (1.00-1.42) | 0.06 | 1 |
| Hyperglycemia | 1.19 (1.03-1.38) | <0.05 | 1 |
| Polypharmacy | 1.23 (1.08-1.41) | <0.05 | 1 |
| Neutrophilia | 1.45 (1.23-1.71) | <0.05 | 1 |
| Elevated CK | 1.70 (1.46-1.99) | <0.05 | 1 |

HR = hazard ratio. CI = confidence interval. Hyperglycemia = glucose > 7 (mmol/L).
Polypharmacy = concurrent medication > 4. Neutrophilia = neutrophil > 7.5 (10<sup>9</sup>/L).
Elevated CK = creatine kinase > 400 (IU/L).

To facilitate practical implementation, we adopted a pragmatic approach by categorizing the suboptimal aspirin treatment effect based on the number of risk factors present in an individual. This risk stratification strategy allows for a straightforward assessment of the likelihood and extent of suboptimal aspirin response based on the number and nature of identified risk factors.

### Model validation

We compared the outcomes in the treatment and control group across different risk categories with different number of risk factors in the validation cohort (Figure 2). For the ease of clinical use and model stability, we defined 3 risk categories regarding the total number of 0, 1-2, and 3-6 risk factors (out of 6) for aspirin treatment effect stratification. Around 80% of patients (79.7% in the treatment group and 78.7% in the control group) were in the risk categories with less than three risk factors. The results showed that patients had similar baseline one-year CVD recurrence risk regardless of the risk category (from 43.5% with 0 risk factors to 52.4% with 3-6 risk factors in the control group). In contrast, the recurrence risk with aspirin treatment showed strong graded association with the risk categories (from 34.0% with 0 risk factors to 54.5% with −6 risk factors in the aspirin treatment group). The stable baseline risk and the graded risk after treatment showed the robustness of the risk factors that we identified. As the number of risk factors increased, there was a gradual approaching tendency in the CVD risk between the treatment and control group. This indicated that aspirin treatment might be more effective in individuals with fewer risk factors than those with multiple risk factors.

**Figure 2.**
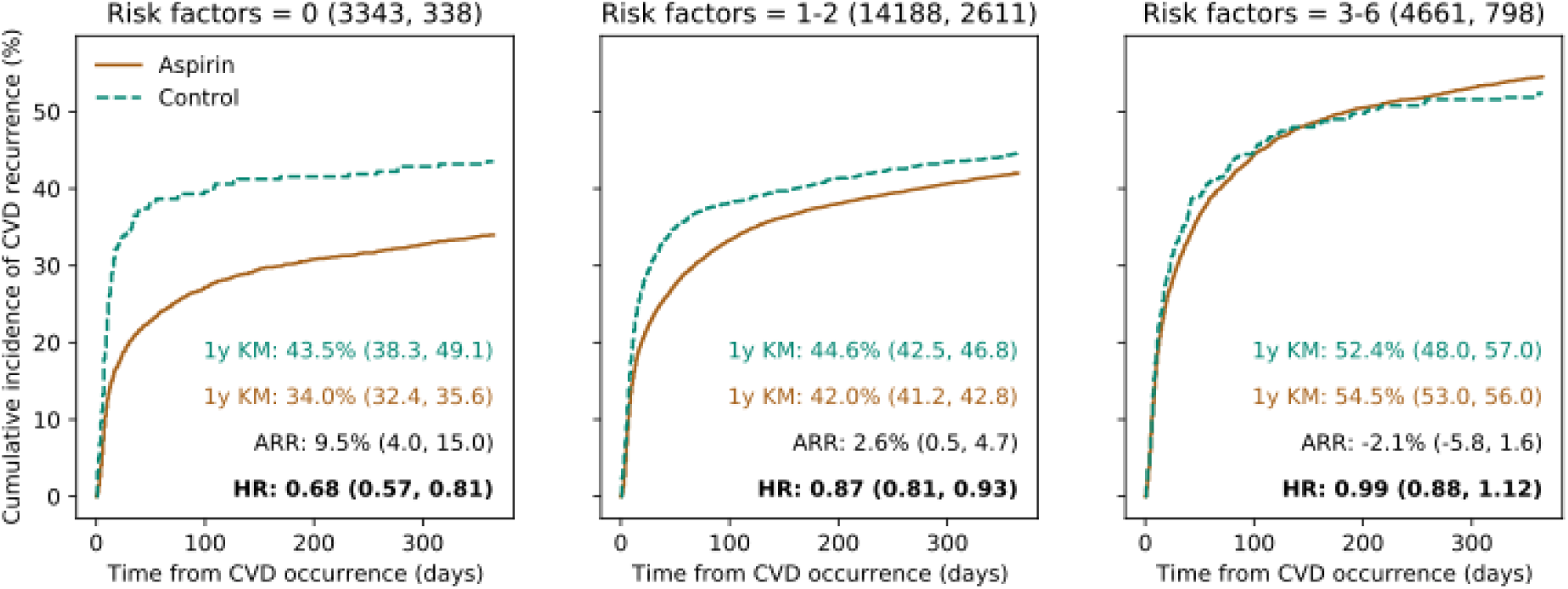
**Outcomes by risk categories and treatment in the validation cohort.** Kaplan-Meier curves stratified by different risk categories and treatment groups (aspirin versus control). Number of patients in aspirin group and control group in each risk category were shown after each subtitle (aspirin, control). One year Kaplan-Meier CVD recurrence probability (1y KM) in the two groups, the corresponding absolute risk reduction (ARR), and the hazard ratio (HR) with 95% confidence interval for aspirin versus control groups are shown.

The HR of aspirin treatment, estimated based on both groups, exhibited a similar trend (Table 4). Specifically, patients with no risk factors (14.2% of patients) had an aspirin treatment HR of 0.68. This suggested that aspirin treatment significantly reduced the risk of CVD events in this low-risk subgroup. For patients with 1-2 risk factors, which constituted 64.8% of the cohort, the aspirin treatment HR is 0.87. Although the effect size was slightly attenuated compared to the no-risk factor group, these individuals still benefited from aspirin treatment as indicated by the reduced HR. In contrast, the remaining 21.0% of patients with 3-6 risk factors exhibited an aspirin treatment HR of 0.99. This group of high-risk patients may be referred to as had aspirin treatment failure, since aspirin therapy did not provide an additional protective effect against CVD events.

**Table 4.** Treatment effect stratification for low-dose aspirin compared to no aspirin treatment after the diagnosis of CVD in the validation cohort (n=25,939)

| Number of<br>VISTA risk factors | One-year aspirin<br>treatment effect (HR) | Number of patients | Proportion of patients |
| --- | --- | --- | --- |
| 0 | 0.677 (0.569-0.806) | 4,403 | 14.2% |
| 1-2 | 0.866 (0.810-0.927) | 16,799 | 64.8% |
| 3-6 | 0.991 (0.876-1.121) | 5459 | 21.0% |

### Website design

A website employed VISTA at www.bio8.cs.hku.hk/vista.html (Supplementary Information 1) was deployed for demonstration, and to enhance utility and clinician-patient discussions.

## Discussion

Our study represents the first investigation, to the best of our knowledge, into the impact of patient characteristics on the effectiveness of low-dose aspirin for preventing recurrent CVD events in secondary prevention. We conducted rigorous statistical testing on a large pool of variables and identified six relevant risk factors, which encompass potential mechanisms underlying suboptimal treatment effects of aspirin. The results of our model validation further demonstrated the strong performance of our model in stratifying aspirin treatment effects. By leveraging VISTA’s capabilities, clinicians gain early access to the potential benefits of aspirin therapy as a treatment option for each patient, allowing them to make informed decisions regarding its suitability. The tool enables clinicians to assess a patient’s risk of treatment failure and proactively adjust the medication regimen by considering alternative therapeutic approaches. For instance, if a patient exhibits a high risk of treatment failure, the clinician can opt to rely on other medications, such as setting lower targets for blood pressure and LDL cholesterol with a higher intensity of antihypertensive and lipid-lowering agents. By optimizing the combination of medications based on individual patient characteristics, this decision support tool has the potential to enhance treatment efficacy and improve outcomes in the secondary prevention of CVD.

Optimizing the prescription of low-dose aspirin for CVD prevention requires a comprehensive evaluation of both the potential bleeding risk and the treatment effect. While a study has attempted on this objective, it only predicted the bleeding risk, without assessing the treatment benefit.^51^ In scenarios where the predicted treatment benefit is unknown, clinicians may only rely on the bleeding risk as a reference. If the bleeding risk is not high, aspirin may be considered as a viable option, even though the benefit remains uncertain. Conversely, if the estimated benefit from aspirin is modest, regardless of how low the bleeding risk is, it is advisable to avoid prescribing aspirin and instead prioritize other medications to achieve the targeted treatment objective. Our study addresses this limitation by directly assessing the treatment effect of aspirin, offering a more comprehensive perspective. Moreover, the bleeding risk study identified 20 risk factors that were associated with the bleeding risk. However, only one of them overlapped with our identified risk factors (diabetes), while the other 19 of them were not associated with treatment failure. This disparity suggests that the treatment benefit and bleeding risk operate through distinct mechanisms and systems. It emphasizes the significance of our study in prompting clinicians to consider the benefit-risk scenario differently when prescribing aspirin.

Aspirin treatment failure has long been recognized as important consideration in clinical practice. Existing methods for assessing aspirin’s antiplatelet effects rely on specific laboratory tests, such as PFA-100, WBA, and VerifyNow, which measure individual platelet function to predict the effectiveness of aspirin. Several case studies have shown that aspirin resistance, as indicated by these assays, is associated with a higher rate of CVD events.^16,52^ However, these tests are not readily available in general hospital settings and have modest correlation and agreement. For instance, in a study of 201 patients with stable coronary artery disease receiving >80mg/day of aspirin, the prevalence of aspirin resistance varied greatly: 7% from VerifyNow, 18% from WBA, and 60% from PFA-100.^18^ Also, they do not directly predict the effectiveness of aspirin in preventing CVD, limiting their practical usefulness for clinicians’ reference. In contrast, our study proposes a practical method that utilizes easily accessible variables to predict the treatment effect of aspirin on CVD prevention. According to the validation results, approximately 80% of patients are expected to experience a considerable treatment effect from aspirin, with hazard ratios ranging from 0.67 to 0.83. However, approximately 20% of patients may have treatment failure (HR close to 1.00) from aspirin if two or more risk factors are present. The findings from our research provide valuable guidance for clinicians in the decision-making process of prescribing aspirin. By offering a straightforward estimation of aspirin’s treatment effect, our method can be used as a tool to assess the potential benefits of aspirin as a treatment option for each patient.

Our work introduced a novel lens to investigate treatment recommendation methods in the era of big data. Traditional approaches to evaluate treatment effects rely on randomized controlled trials (RCTs). RCTs face practical constraints, cost considerations, and data management complexities, which often lead to the collection of a limited number of patient characteristics. While RCTs are reliable in yielding population-level outcomes, stratifying individual benefits based on these limited covariates is challenging. In our research, we demonstrated the potential of high-quality retrospective cohort studies with a comprehensive set of covariates in treatment effect stratification. We utilized a robust feature selection process to identify key risk factors in aspirin efficacy variation and applied rigorous validation to estimate the variation of the treatment effects across various risk groups. In the era of big data and with advances in data curation, there is an opportunity to develop more robust treatment effect stratification that complements the population-level results obtained from RCTs. Our approach has proved to be robust in addressing this task and can be extended to analyze the efficacy of various therapies. By leveraging the vast amount of available data, we can gain valuable insights into individual treatment responses and enhance personalized medicine approaches.

There are limitations in our study. First, both our derivation and validation cohorts were developed using data from Hong Kong. Although Hong Kong is a multi-ethnic city, its population is predominantly Chinese. Consequently, the generalizability of our model’s performance to other ethnicities and regions may be uncertain. Future research should aim to apply our model to validation cohorts comprising diverse ethnic populations to assess its performance across different racial groups. Second, our estimation of treatment effects across risk groups was based on a retrospective cohort study design. This introduces the possibility of selection bias in the selection of the treatment and control groups. To further validate our model, it would be beneficial to conduct more rigorous investigations, such as prospective clinical studies that incorporate the collection of the identified key risk factors. This independent validation of each risk factor and evaluation of the performance of risk group treatment stratification would provide stronger evidence for the effectiveness and reliability of our model.

## Conclusions

The variability in aspirin treatment effects poses a significant challenge in optimizing therapeutic outcomes for patients with CVD. VISTA can be helpful in the secondary prevention of CVD which can identify patients who may be at risk of treatment failure thus help clinicians with treatment planning. We identified six key risk factors, history of atrial fibrillation, dyslipidemia, hyperglycemia, polypharmacy, neutrophilia, and elevated CK, that contribute to the variation in aspirin treatment effects. The big-data approach employed in our study also holds promise across various therapeutic areas beyond aspirin therapy. This research represents a significant step towards advancing personalized medicine in aspirin therapy, ultimately enhancing therapeutic effectiveness and improving CVD prevention on an individualized basis.

## Data Availability

Data will not be available for others as the data custodians have not given permission.

## Acknowledgements

Nil.

